# Scientific output on coronavirus and WHO’s Solidarity Project: a science-based choice?

**DOI:** 10.1101/2020.11.16.20232488

**Authors:** Andréia Cristina Galina, Deise Sarzi, Larissa Campos de Medeiros, André Luiz Franco Sampaio, Jacqueline Leta

## Abstract

In March 2020, the World Health Organization (WHO) launched the Solidarity Program probably the largest global initiative to encourage and support research in four promising drugs and therapies (Remdesivir, Hydroxychloroquine, β interferon and the combination Lopinavir / Ritonavir) to reduce the mortality of COVID-19. Considering the potential impact of this project to restrain the current pandemic, the present study aims to investigate whether it was designed upon a scientific basis. For this proposal, we collected all documents on coronavirus indexed in Scopus database by using a search strategy based in MESH terms. Among the studied groups of documents, we looked in more detail the Coronavirus group in order to find documents related to WHO’ s drugs or to other drugs and therapies extracted from another source. The main findings indicate that the number of documents related to WHO’s drugs are higher than in the other groups and this subset of documents involves a larger number of institutions and countries. Hence, the results shown in this study illustrate that decisions by an international body, as WHO, may be science-based and not be merely bureaucratic decisions.

## INTRODUCTION

Although coronaviruses became popular in 2020, these types of viruses have been considered pathogens for several groups of animals, including humans, since decades. Today, it is widely known that coronaviruses are a large family of viruses with the potential to cause a spectrum of diseases ranging from a simple cold to a severe acute respiratory syndrome (1).

In the 1960s, coronaviruses that infect humans were isolated for the first time from samples of nasal secretion of patients with symptoms of cold (2). Since then, seven types of human coronavirus (HCoV) have been identified, among which four are endemic, that is, are exclusively found in some regions and are associated with mild symptoms of respiratory diseases (HCoV-229E, HCoV-OC43, HCoV-NL63 and HCoV-HKU1). Two other types include the acute respiratory syndrome virus (SARS-CoV), identified in 2002, and the Middle East syndrome virus (MERS-CoV), identified in 2012, both are associated with severe respiratory symptoms and high mortality rates (3). The last type, identified in 2019 and known as “new coronavirus” (SARS-CoV-2), is also associated with severe respiratory symptoms, but unlike previous ones, it is highly transmissible (4; 5), which favored the rapid spreading of the current pandemic.

According to the National Center for Biotechnology Information (NCBI), coronaviruses belong to the family Coronaviridae, subfamily Orthocoronavirinae, which is divided into four genera: Alphacoronavirus, Betacoronavirus, Gammacoronavirus and Deltacoronavirus. Alpha and Betacoronavirus infect only mammals, while most gamma and Deltacoronavirus infect birds but it may also infect mammals (6; 7). Out of the seven HCoV already identified, two belong to the genus Alphacoronavirus (HCov-229E and HCoV-NL63) and the other five belong to the genus Betacoronavirus (7).

The Betacoronavirus SARS-CoV-2, that is, the new coronavirus is highly pathogenic and responsible for the coronavirus disease pandemic started in late 2019, named COVID-19. This pandemic has been mobilizing the global scientific community in the search for alternatives to diagnose and prevent new cases, as well as alternatives to treat patients already infected by SARS-CoV-2 (8-10). The current efforts of scientists from different parts of the world to increase and to share rapidly the new knowledge on COVID or coronavirus have been discussed and presented in several studies (11-15).

The growing literature about scientific publications on COVID or coronavirus at a time when the world still suffers from the pandemic has highlighted not only general analyzes such as the studies mentioned above, but has also emphasized other aspects including: the contribution of a specific country or region (16, 17), comparison with the scientific publication on other viruses (18), publication analysis based in different information sources (19, 20) and about the faster editorial flow (21). Despite the wide thematic variety in this literature, we have not observed studies focusing on scientific publication on drugs and preventive treatments, such as vaccines, to restrain the new coronavirus.

The discovery and development of new drugs, as well as other treatments, can take years to result in a prototype and product (10, 22, 23). However, in an emergency situation, one way to fasten this process is the repositioning of drugs, that is, the investigation of a new use for an existing drug, thus avoiding expensive and time-consuming toxicological essays (24, 25). Such initiatives have been taking place in the COVID-19 epidemic, being the Solidarity Program probably the largest example. Launched in March 2020 by the World Health Organization (WHO), the program aims encouraging and supporting worldwide research in four promising drugs and therapies (26, 27): (i) remdesivir, an antiviral that inhibits the RNA virus cycle, (ii) hydroxychloroquine, a drug used as a therapy against malaria and rheumatoid arthritis, (iii) berta interferon, a drug used in the treatment of multiple sclerosis and (iv) the combination lopinavir / ritonavir, drugs used to treat HIV. After almost four months and in the face of negative results, WHO reviewed the program and withdrew the support for research with hydroxychloroquine and the combined drugs lopinavir/ritonavir.

Although the motivation for choosing these therapies, and not others, is not clear in the Solidarity Project (20), it is expected that the WHO’s choice was based on existing scientific evidences that place these therapies in a prominent position. However, we may not discard that many national and even global initiatives, in this case related to the epidemic, can also be motivated by other aspects, such as politics, as was observed in a recent study (28).

Considering both the lack of studies about research on drugs related to coronavirus and the potential impact of the Solidarity Project in to restrain the current pandemic, the present study aims to investigate whether WHO’s project was designed upon a scientific basis, that is, the extent to which the drugs indicated in the project reflect the efforts of the global scientific community related with coronavirus. For this, we carried out a comparative bibliometric study regarding both the drugs indicated in the Solidarity Project and other drugs and vaccines already in an advanced stage in the clinical trials, as indicated in the Report “Disease Briefing: Coronaviruses”, by Clarivate Analytics (29). The Clarivate report in March 2020, by the time we started collecting data, was an international reference with wide coverage and details on coronavirus research, compiling information from Cortellis, Web of Science, and BioWorld.

## MATERIALS AND METHODS

We used the Medical Subject Headings (MeSH) browser (30) to define the search strategy. On April 25, 2020, by using the exact match term ‘coronavirus’ in the filters Full Word Search, Exact Match and Main Heading Terms, we found three main terms in MeSH Tree Structures, which we named *coronavirus groups*, as presented in Figure 1. Note that the Deltacoronavirus is not presented among the three main groups in the MeSH Tree Structure, which explains its absence in the following analysis.

**Figure 1:**
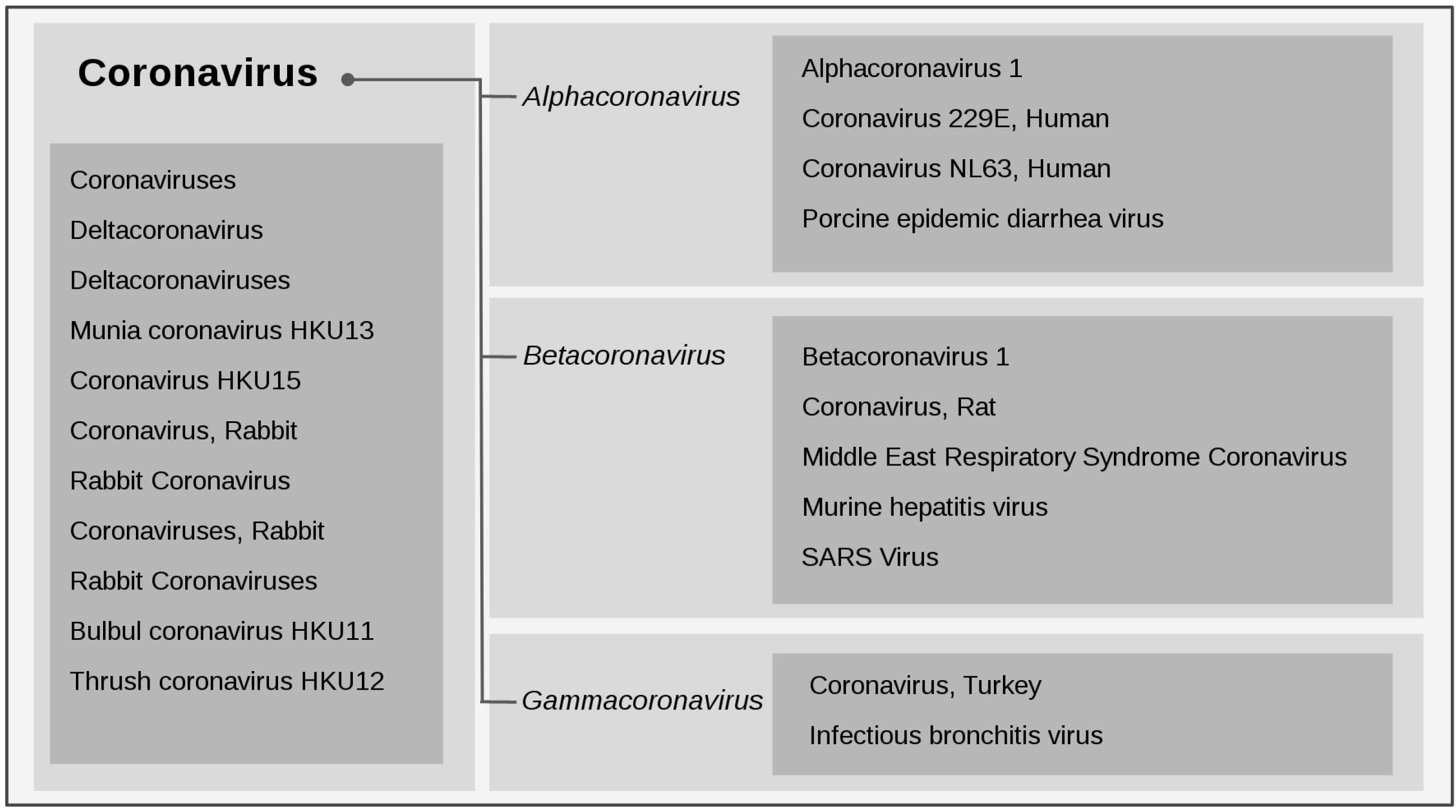
MeSH Tree Structure based in the exact match term coronavirus.

After testing variations of MeSH terms, we decided to collect data separately in four groups: a generalist and other three groups that correspond to coronavirus genera found in MeSH Tree. The search strategies used to collect documents for each group are as following:

- Coronavirus: coronav*;
- Group Alphacoronavirus: alphacoronav* OR “alpha coronav*” OR alphacoronavirus-1 OR “Transmissible gastroenteritis vir*” OR hcov-nl63 OR hcov-229e OR “Porcine epidemic diarrhea vir*”;
- Group Betacoronavirus betacoronavirus-1 OR betacoronav* OR beta-coronav* OR hcov-hku1 OR (“Porcine hemagglutinat*” W/1 “encephalomyelitis vir*”) OR “MERS vir*” OR mers-cov OR (murine W/1 “hepatitis vir*”) OR (mouse W/1 “Hepatitis Vir*”) OR (“Gastroenteritis Vir*” W/1 murine) OR mhv-jhm OR “SARS Vir*” OR (“Severe Acute Respiratory Syndrome” w/1 vir*) OR sars-cov*.
- Group Gammacoronavirus: gammacoronav* OR “Gamma coronav*” OR “Bluecomb Vir*” OR (“Transmissible Enteritis Vir*” W/2 turkey*) OR (“Enteri* Vir*” w/3 Turkey) OR (“bronchit* vir*” W/1 infect*);

Data collection was carried out on May 30, 2020 at the Scopus database, due to its greater collection coverage when compared to other databases. Using the advanced search mode, the search strategy of each group was searched in title, abstract and keyword filters. The results were exported in the BibTeX format considering data on Citation information, Bibliographical information, Abstract & keywords and Funding. The totals of retrieved documents in the four groups were: 2,719 documents on alphacoronavirus, 13,655 on Betacoronavirus, 2,745 on Gammacoronavirus and 28,013 on coronavirus. The later, that is, the most generalist and largest group, was used as reference for all analysis, with the exception of Figure 2.

**Figure 2:**
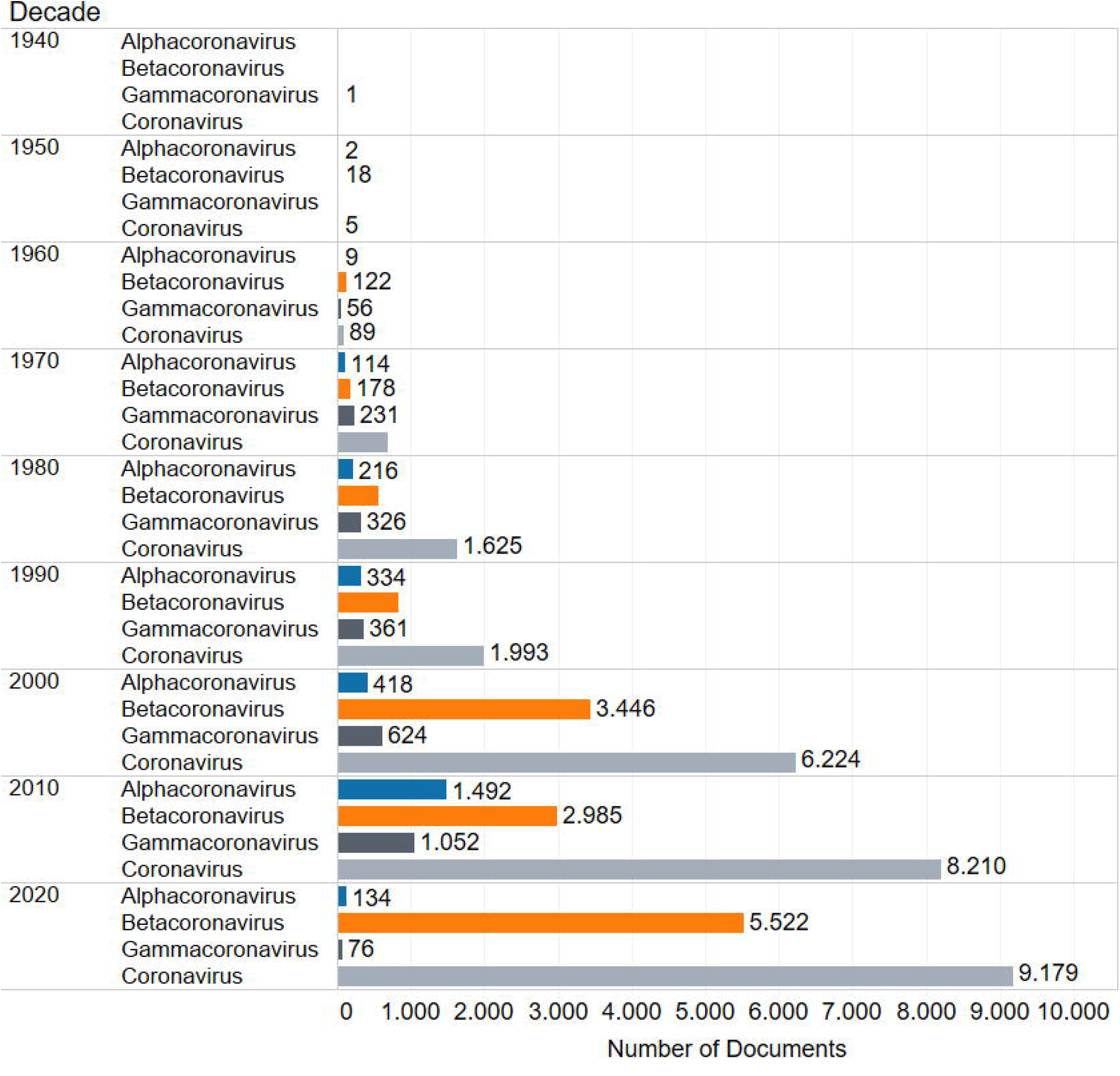
**Number of documents on coronavirus and on the three coronavirus families according to the decade. Source: Scopus**

The 28,013 documents on coronavirus were classified according to the type of research application as: vaccine, diagnosis and treatment. To identify documents classified as vaccine, we used the words *vaccine, vaccines, vaccinia* and *vaccination*. As for diagnosis, we used the *diagnos* while for those classified as treatment, we used the words *treat* and *therap*. For this process, we used a script developed in the R language (31) and the words were searched in the title, abstract and database and author’s keywords.

Documents on coronavirus were also classified according to the drug or therapy they are related with, that is, WHO’s drugs and other drugs or therapies listed in the Clarivate’s report. To identify documents that are related with WHO’s drugs, we used the following terms: “hydroxychloroquine sulfate”, remdesivir, “lopinavir / ritonavir”, “ritonavir / lopinavir”, “lopinavir plus ritonavir”, “ritonavir plus lopinavir”, “lopinavir ritonavir”, “ritonavir lopinavir”, “interferon beta”, “beta interferon”, “interferonbeta” and “betainterferon”. As for second group, initially we have selected 31 drugs or therapies all listed as clinical trials in phases I, II or III in Clarivate’s report. Nevertheless, we discarded 11 drugs and therapies since they were not found within documents on coronavirus. Thus, documents related to 20 drugs and therapies were classified in three groups: other drugs, antibody and vaccine. The group Other drugs includes documents with at least one of the terms: “darunavir/cobicistat”, “cobicistat/darunavir”, “darunavir plus cobicistat”, cobicistat plus darunavir”, “darunavir cobicistat”, “cobicistat darunavir”, “chloroquine phosphate”, “danoprevir”, “diammonium glycyrrhiinate”, “favipiravir”, “leronlimab” or “oseltamivir phosphate”. As for the group Antibody, documents include at least one of the terms: “sarilumab”, “regeneron 3048”, “regn 3048”, “regn3048”, “regn-3048”, “regeneron 3051” “regn 3051”, “regn3051”, “regn-3051”, “sab-301”, “sab 301”, “sab301” or “siltuximab”. Finally, the group *vaccine* includes documents with at least one of the following terms: “gls5300”, “gls-5300”, “gls 5300”, “chadox1 mers”, “chadox1-mers”, “mva-mers-s”, “mvamers-s”, “mrna 1273” or “mrna-1273”. All these terms were searched in the abstract, document title and database and author keywords.

For the analysis of institutions, we have considered the data on “author’s affiliations – disambiguated” of documents classified as WHO’s drugs or Other drugs, Antibody and Vaccine. With the help of a script developed in R language, affiliations were checked and duplications in a single document were deleted. Thus, based on the list of institutions without duplications, it was possible to rank institutions and countries according to the frequency of documents in each group.

In order to verify whether the most prolific institutions among documents on coronavirus are also the most prolific in terms of the development of therapies against COVID-19, in 23 July, 2020, we collected the list of agreements in progress catalogued by Cortellis, a database of Clarivate Analytics. By searching the term “coronavirus infection”, we have found 2,226 agreements, which encompass 186 institutions, 766 drugs and 879 patents. Data on agreements were downloaded in a Microsoft Excel format and all agreements dealing with drugs and therapies were identified as following.

Agreements classified as WHO’s drugs were identified by having the following terms: hydroxych, remdesivir, lopinavir, ritonavir, “interferon beta”, “beta interferon”, interferonbeta and betainterferon. As for agreements on Other drugs, we used the words darunavir, cobicistat, chloroquine, danoprevir, diammonium, glycyrrhizinate, favipiravir, leronlimab and oseltamivir. As for agreements classified as antibody and vaccine were identified, respectively, by containing at least one of the terms (a) sarilumab, “regeneron 3048”, “regn 3048”, regn3048, regn-3048, “regeneron 3051”, “regn 3051”, regn3051, regn-3051, sab-301, “sab 301”, sab301 and siltuximab and (b) gls5300, gls-5300, “gls 5300”, chadox1, mva, “mrna 1273” and mrna-1273. The affiliations of each group of agreement were identified and the frequency of institutions and countries was then calculated.

## RESULTS

The sum of the documents classified in all four studied groups (Alphacoronavirus, Betacoronavirus, Gammacoronavirus and Coronavirus) with no duplication is 31,815. This amount is distributed in the following typologies: 68% article or article in press, 10% article review, 7% letter, 5% editorial, 4% note, 3% conference paper or review, 1% book, 1% short survey and 1% data paper, erratum or retracted. In the following sections, we present the results regarding (a) general data, including the time trends of documents on each group of coronaviruses and the number of documents on Coronavirus group according to the type of research application and (b) a subset of data on Coronavirus groups related with drugs and other therapies as well as their leading institutions and countries.

### Time trends of the four groups of documents on coronavirus

The number of documents on coronavirus classified under the four main groups by decades is shown in Figure 2. As can be seen, the number of documents found in the 1940s and 1950s is residual, either because of the database’s low journal coverage in these decades or because research on coronavirus was still incipient at that time.

In the 1940s, we found the first document on coronavirus registered in Scopus: a paper entitled “Demonstration of an interference phenomenon associated with infectious bronchitis virus (IBV) of chickens”, that is, a clear example of document related to one of the main MeSH terms related to Gammacoronavirus. As for the following decade, we found 18 documents in the Betacoronavirus group, being none of them related to human coronavirus but to hepatitis virus either in mouse or mice, which is in accordance to MeSH terms indicated to this family.

From 1960s to 2020s, there was a strong growth trend in the number of documents related to most groups, specially the Coronavirus group (gray bar). Documents of this group increased from 89 to 9,179, that is, a growth rate of 102.1 in the period. Considering the total of documents indexed at Scopus database, these totals represent 0.00% and 0.44%, respectively, which signals a real increase in this theme in the set of documents in the database.

The total of documents related to the three coronavirus genera also indicates growth over the periods, except for 2020 in the groups of Alphacoronavirus (blue bar) and Gamacoronavirus (black bar), being the latter not associated with humans. The total number of documents in these two groups increased, respectively, from 9 to 1,492 (a growth rate of 164.8) and 56 to 1,052 (a growth rate of 17.8) in the period from 1960 to 2010. Considering the whole set of coronavirus documents (with no duplication) published in the 2010s (n = 8.210), the share of documents on Alphacoronavirus and Gamacoronavirus are 18.17% and 12.81% respectively.

As for Betacoronavirus (orange bar), we observe an intense growth until 2020 with an increased from 122 in 1960 to 5,522 in 2020 (a growth rate of 44.3). These totals correspond to 0.38% and 17.35 % of the whole set of coronavirus documents (with no duplication) published in the respective decades. It is noteworthy that, as observed for the coronavirus group (gray bar), documents on Betacoronavirus had a notable increase over the period, which seems to take place in two phases: from 1960 to 1990 and from 2000 to 2020. In this second phase, we observe a greater acceleration in the growth rate. The inclusion of new titles in the Scopus database throughout the 2000s may partially explain it while the outbreak of SARS in Asia during 2020, which reached a total of 33 countries on the five continents (32), has boosted research on coronavirus, especially on Betacoronavirus that is associated with SARS, and so led to the notable increase in publications observed between the 1990s and 2000s.

### Types of research application among documents on coronavirus group

As the Coronoravirus group (n. 28,013) display the largest number of documents, we decided to investigate some parameters to better characterize it. In this section, we present the distribution of these documents according to the type of research application they are related with, that is (a) diagnosis, (b) treatment or (c) vaccine, a typology is supported by Tarik Jasarevic (33). The analysis was carried out considering a Venn diagram (Figure 3), which makes it possible to identify the intersections between the three types of applications.

**Figure 3:**
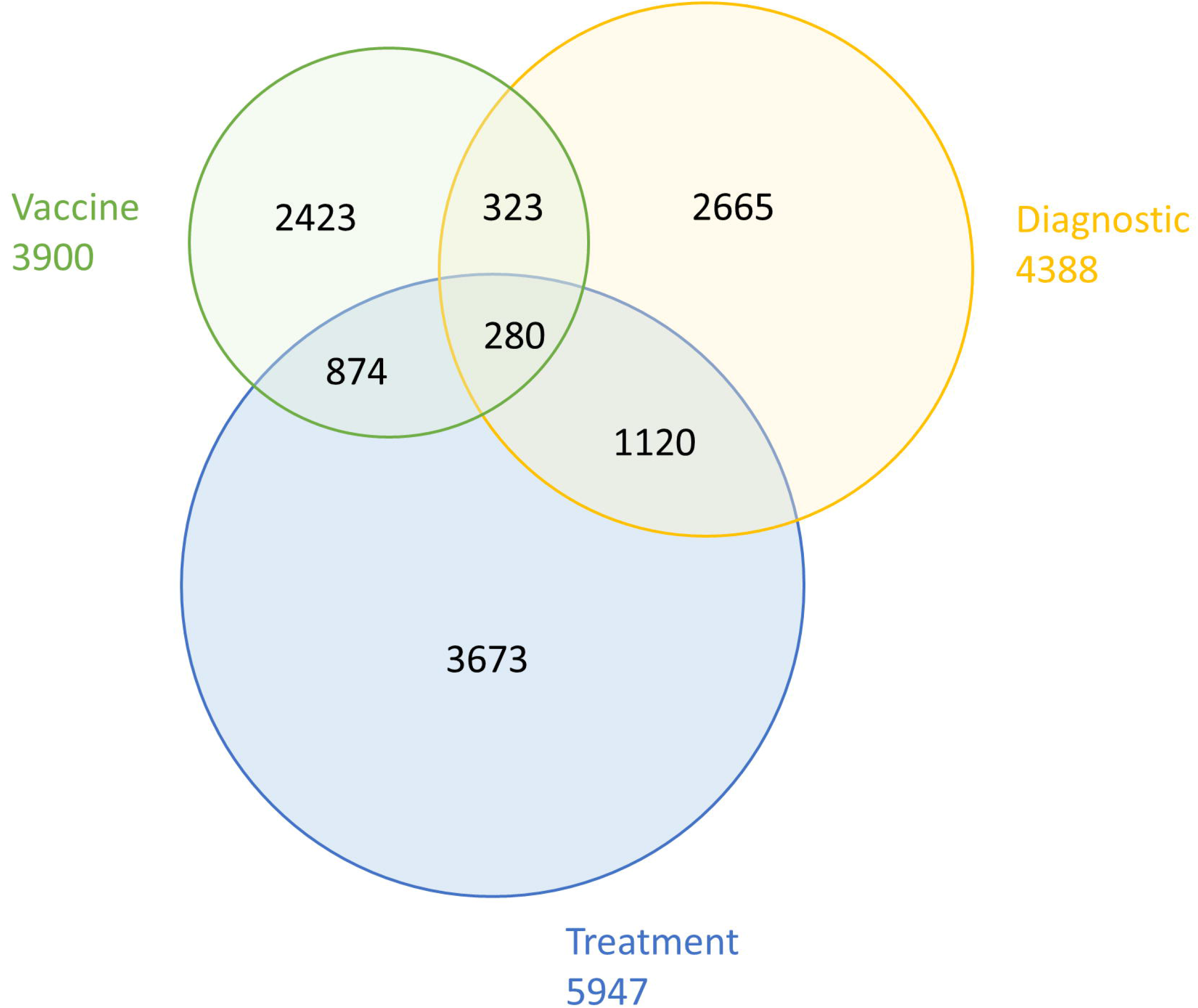
**Venn diagram to the three types of research application among documents on coronavirus published in the complete period. Source: Scopus**

We found that, over the period of analysis, the scientific community on coronavirus has been dedicating more efforts to research related to treatment (n. 5,947). The documents in this group, in general, include controlled studies with antiviral agents or with some other drugs to reduce or combat symptoms related to the disease.

Documents related to diagnosis (n. 4,388) and vaccine (n. 3,900) are less frequent. The first group includes publications that deal with diagnostic imaging techniques, techniques for the detection of the virus and epidemiological data[, while the latter includes publications related to techniques and essential content for the development of a vaccine, such as the viral genome, the viral envelope proteins, neutralizing antibodies, virus replication etc.

Regarding the shared areas in the diagram, documents related to treatment display the largest intersection areas, they are: with diagnosis (n. 1,400 documents) and with vaccine (n. 1,154 documents). This finding reinforces the prominent role of this type of research application (treatment) within the set of documents on coronavirus. It is also relevant to highlight the 280 documents that were identified in the intersection area that includes treatment, diagnosis and vaccine. Out of 280 documents, 130 are review articles, while the remaining are research articles focused in a more general understanding of the disease or the virus.

### Drugs and other therapies among documents on coronavirus group

In this section, we identify and compare the number of documents on Coronavirus group (n. 28,013) related to two groups of drugs and to a group of Antibody, both with the potential for use in the treatment of COVID-19 patients. We also analyzed the documents on coronavirus related to a group of vaccines, which makes a counterpoint to the groups of drugs and Antibody, since it is generally a preventive method for avoid spreading the disease. Figure 4 presents the number of documents on coronavirus by decade according to the group of drugs or other therapies. As a first observation, we draw attention to the low number of documents in the four groups, which adds up to 908 between 1980 and 2020, that is, a share of 3.24% of the total documents on Coronavirus group published in the period.

**Figure 4:**
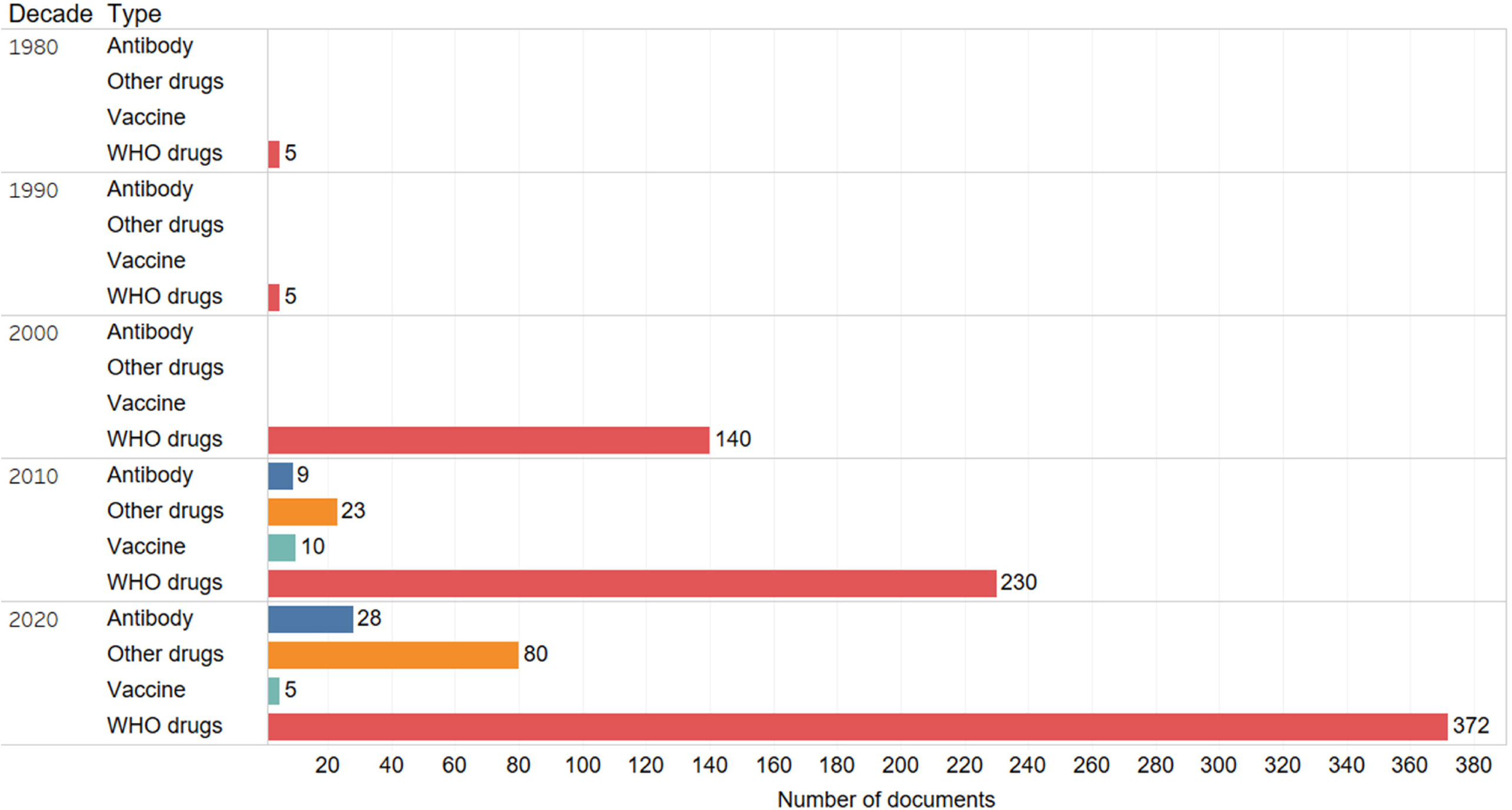
**Number of documents on coronavirus main groups related to drugs and other therapies by decade. Source: Scopus**

As can be noted, WHO drug-related documents (red bar), that is, those indicated in the WHO Solidarity Project, predominate largely among the documents on coronavirus in all periods. Documents included in WHO drugs sum 752, that is, 82.8% of the total number of documents on coronavirus included in the four groups of drugs or other therapies.

From the 2010s on, it is noted an increase in documents related to other drugs (orange bar) and antibody (dark blue). We believe that such increase is due to the outbreak of MERS-CoV, which emerged in Saudi Arabia in 2012. The research driven by this outbreak gained more strength in late 2019, when the SARS-CoV2 pandemic started.

Curiously, we did not observe a similar trend among documents related to vaccines (light blue). The low and reducing number of these documents in the last decades may be a consequence of the complex process that developing a vaccine is, characterized by its time duration and confidentiality, which often reduces the diffusion of the publication.

For a better understanding of the nature of the four groups of drugs and other therapies, Table 1 presents the name of the compound and its respective number of documents in each group.

**Table 1:** **Compounds and their respective number of documents on coronavirus main groups related to drugs and other therapies, 1940 to 2020. Source: Scopus**

| Drugs / Sources | WHO |  | Clarivate |  |
| --- | --- | --- | --- | --- |
|  | Name | N. | Name | N |
| Drugs /<br>other drugs | Hydroxychloroquine sulfate | 13 | Chloroquine Phosphate | 5 |
|  | Interferon Beta | 331 | Danoprevir | 5 |
|  | Lopinavir/ritonavir | 325 | Darunavir/cobicistat | 16 |
|  | Remdesivir | 257 | Diammonium glycyrrhizinate | 1 |
|  |  |  | Favipiravir | 88 |
|  |  |  | Leronlimab | 2 |
|  |  |  | Oseltamivir phosphate | 1 |
| Antibody |  |  | REGN 3048 | 4 |
|  |  |  | REGN 3051 | 3 |
|  |  |  | SAB-301 | 6 |
|  |  |  | Sarilumab | 24 |
|  |  |  | Siltuximab | 9 |
| Vaccine |  |  | ChAdOx1 MERS | 3 |
|  |  |  | GLS 5300 | 5 |
|  |  |  | mRNA 1273 | 4 |
|  |  |  | MVA-MERS-S | 3 |

Within the group WHO’s drugs, documents referring to a drug named Interferon beta are the majority (n. 331), while those related to Hydroxychloroquine sulfate, which was widely propagated by some global representatives (34) as a potential treatment for COVID 19, are the least frequent (n. 13) in this group.

Regarding the group Other drugs, we found documents on coronavirus related to seven drugs, among them Favipiravir, a nucleoside analog already used in the treatment of influenza A and B, appears with the largest number of documents in the period (n. 88). In the group Antibody, we identified documents on coronavirus related to five drugs, being Sarilumab, a human monoclonal antibody already used to treat rheumatoid arthritis, the one with the largest number of documents (n. 24). Finally, in the group Vaccine, the largest number of documents is related to the GLS 5300 vaccine (n. 5), a vaccine that uses plasmid DNA to express the MERS-CoV spike glycoprotein.

### Drugs and other therapies among coronavirus documents and agreements: the main leaders

In this final section, we analyze the institutions and countries that lead the research in the four groups of drugs and other therapies according to the number of documents on Coronavirus group (collected from Scopus database) as well as agreements to develop therapies against the coronavirus (collected from the Cortellis database).

In Table 2, we first present a general analysis, where the number of documents and agreements on coronavirus are related to the number of institutions and countries within the four main groups of drugs and other therapies.

**Table 2:** **Number of documents on coronavirus and active agreements on coronavirus infection and respective number of institutions and countries according to the four main groups of drugs and other therapies**.

| **Variable / Sources** | **Documents from Scopus** |  |  |  | **Agreements from Cortellis** |  |  |  |
| --- | --- | --- | --- | --- | --- | --- | --- | --- |
|  | *WHO drugs* | *Other drugs* | *Antibody* | *Vaccine* | *WHO drugs* | *Other drugs* | *Antibody* | *Vaccine* |
| ***N. Documents / Agreements*** |  |  |  |  |  |  |  |  |
| With 1 institution address | 270 | 34 | 12 | 2 |  |  |  |  |
| With 2 institution addresses | 216 | 35 | 8 | 3 | 39 | 47 | 15 | 30 |
| With 3- 6 institution addresses | 266 | 35 | 17 | 9 |  |  |  |  |
| ***N. Institutions*** | 1,310 | 260 | 152 | 38 | 52 | 62 | 15 | 33 |
| ***N. Countries*** | 67 | 33 | 26 | 11 | 13 | 19 | 6 | 12 |

Source: Scopus (May, 2020) and Cortellis (July, 2020)

In Scopus documents, we observed that documents in WHO drugs and Other drugs are distributed almost equally among the three categories of number institutions. Documents with two or more institutions sum 482 and 70, which represent 64.1% and 67.3% in both groups, respectively. As for documents in Antibody and Vaccine, we note a quite different trend, especially the latter, with 80% of the documents signed by two or more institutions. Although documents of this groups tend to show a high level of collaboration, they embrace the lowest diversity and number of institutions and countries.

As for the agreements on coronavirus, that is the Cortellis dataset, we did not observe any variation in terms of number of institutions, since all agreements refer the participation of two institutions only, in other words, data refer to bilateral agreements. However, we noted a low level of institutional and country diversity, especially among documents included in Antibody and Vaccine groups. When compared to Scopus dataset, we observed a different trend in terms of the share of each group: the largest number of agreements is related to Others drugs rather than to WHO’s drugs while the lowest number of agreements is related to Antibody and not to Vaccine.

In order to better understanding whether institutions are involved in the research on coronavirus and in the development of a treatment to prevent or to seek the cure for the virus, we investigated the top leading institutions in terms of number of documents and agreements according to the groups of drugs and therapies (Table 3).

**Table 3:** **Top ranked institutions affiliated to documents on coronavirus and to agreements on COVID-19 according to the four main groups of treatment**.

| Drugs / Sources | Documents from Scopus | Agreements from Cortellis |
| --- | --- | --- |
| WHO drugs | University of Hong Kong (47), University of North Carolina (24), Vanderbilt University (19), University of Toronto (14), University of Pennsylvania (13), Harvard Medical School (12), University of Texas (12), University of Virginia (12), University of Iowa (11) and Cleveland Clinic Foundation (10) | Gilead Sciences Inc (13), Abbott Laboratories (4), AbbVie Inc (3), Bayer Schering Pharma AG (3), Synairgen plc (2), Accord Healthcare Inc (2), National Institute of Allergy and Infectious Diseases (2), Novartis AG (2), Triangle Pharmaceuticals Inc (2), University of Oxford (2) |
| Other drugs | University of Hong Kong (6), Karolinska Institutet (4), Peking University (4), National Taiwan University Hospital (3) e University of Virginia (3), Academic Medical Center (2), Aix-Marseille Universit (2), Al-Faisal University (2), Emory University (2) e Fudan University (2). | CytoDyn Inc (15), Drexel University College of Medicine(3), FUJIFILM Toyama Chemical Co Ltd (3), InterMune Inc (3), Progenics Pharmaceuticals Inc(3), Ajinomoto Althea Inc (2), Ascleptis Pharma Inc (2), FUJIFILM Holdings Corp (2), Janssen Diagnostics BVBA (2), Johnson & Johnson (2), MediVector Inc (2), National Institute of Allergy and Infectious Diseases (2), Roche Holding AG (2), Russian Direct Investment Fund (2), Toyama Chemical Co Ltd (2) |
| Antibody | Columbia University (4), Istituto Nazionale Tumori (3), Harvard Medical School (3), National Institute Of Allergy and Infectious Diseases (3), MacroGenics Inc. (2), Yale University School Of Medicine (3), Sapienza University Of Rome (2), Johns Hopkins University School Of Medicine (2), National Institutes Of Health (2) e Regeneron Pharmaceuticals (2) | Regeneron Pharmaceuticals Inc (8), EUSA Pharma (4), US Department of Health and Human Services (4), SAB Biotherapeutics Inc (2), Sanofi SA (2) |
| Vaccine | German Center For Infection Research (4), Philipps University Of Marburg (4), University of Munich (3) and University of Oxford (3) | ModeRNA Therapeutics (9), AstraZeneca plc (6), The Jenner Institute (6), University of Oxford (6), Coalition for Epidemic Preparedness Innovations (4), US Department of Health and Human Services (2) |

Among Scopus documents, we observed the predominance of institutions from education sector and the presence of a few number of research institutes and corporations. In the two groups related to drugs (WHO drugs and Other drugs), the University of Hong Kong appears with the largest number of documents. Other universities also stand out in these groups, but are not listed in the others groups, they are: Hong Kong University, Emory University and University of Virginia. Out of the institutions that are listed in more than one group, apart those mentioned before, we found Harvard Medical School and University of Iowa, both authoring documents on WHO drugs and Antibody.

In the group Antibody, the leadership is with Columbia University. This group presents the largest variety of research institutes and corporations, such as the Istituto Nazionale Tumori, National Institute of Allergy and Infectious Diseases, Macrogenics Inc., National Institutes of Health and Regeneron Pharmaceuticals. While the group Vaccine shows German Center for Infection Research and Philipps University of Marburg (both assigning four documents) and other two universities, University of Munich and University of Oxford. None of the institutions in this group is repeated in the others.

A comparison with the top leading institutions in terms of agreements on coronavirus pointed to a completely different profile, with a predominance of private companies established in Europe and the USA, mainly. We also observed the presence of a few number of universities and research institutes, of which we stand out: (a) The National Institute of Allergy and Infectious Diseases (USA) included in the groups WHO’s drugs and Other Drugs, (b) the University of Oxford (UK) enclosed in the groups WHO’s drugs and Vaccine and (c) the US Department of Health and Human Services (US) in the groups Antibody and Vaccine.

Among the private companies, we highlight the following: (a) in the group WHO’s drugs, the Gilead Sciences Inc that is investigating the drug Remdesevir (35), (b) in the group Other Drugs, the CytoDyn Inc that is carrying on research on Leronlimab which is in phase 3 clinical trial (36) and (c) in the group Vaccine, the ModeRNA Therapeutics that is developing the mRNA-1273 vaccine, which is also in phase 3 clinical trial (37).

As a final analysis, we investigated the distribution of documents and agreements on coronavirus according the country, as shown in Figures 5 and 6. The color intensity in both maps indicates the quantity of documents or agreements in each group of drugs or therapies. To better visualize the contribution of each country, the maps present an attached table, which contains the total number of documents (Figure 5) or agreements (Figure 6) by country as well as the number of documents or agreements in collaboration (internal collaboration, that is, between institutions in the same country, and external collaboration, that is, between institutions from different countries).

**Figure 5:**
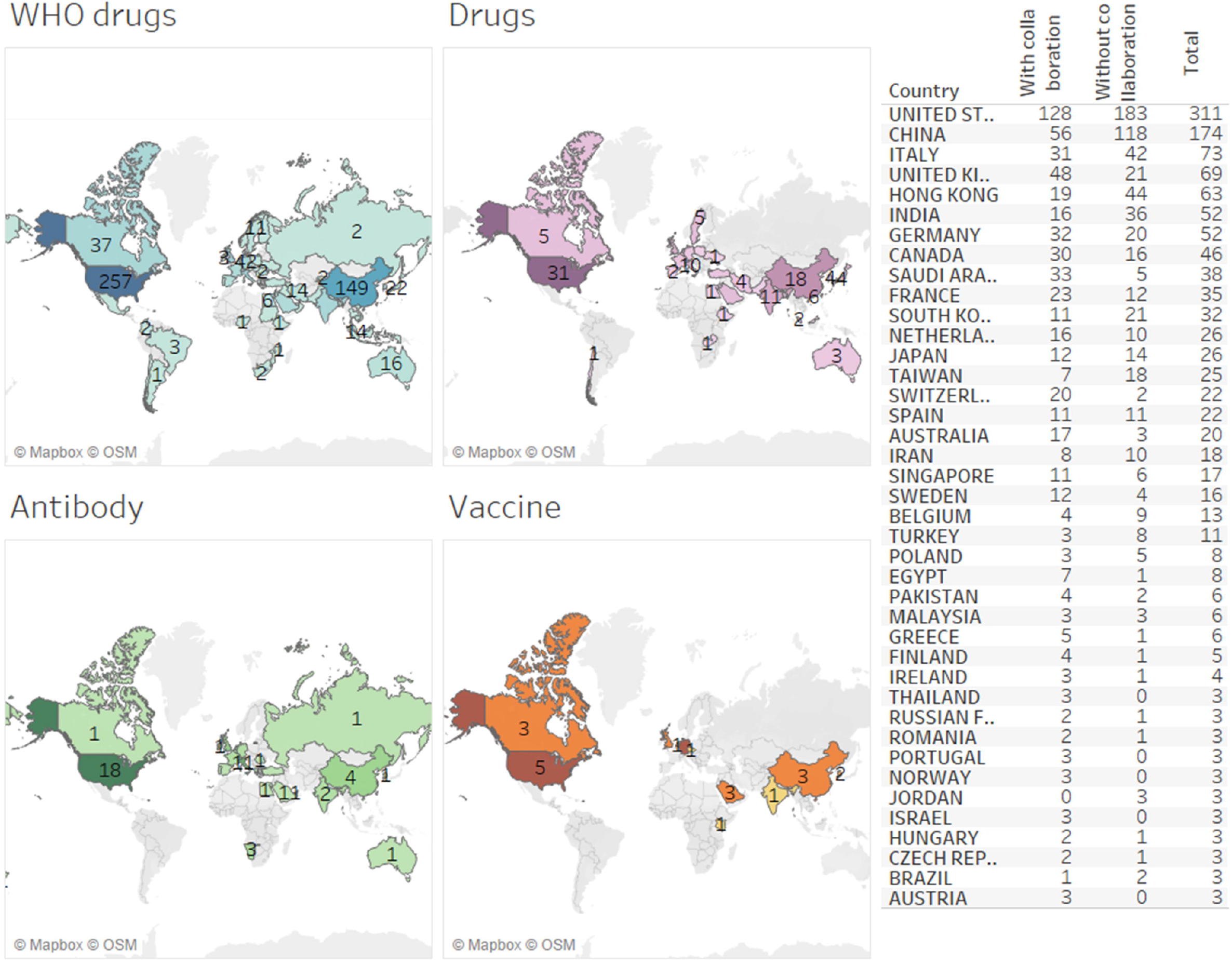
**Documents on coronavirus per country according to the four main groups of treatment. Source: Scopus**

**Figure 6:**
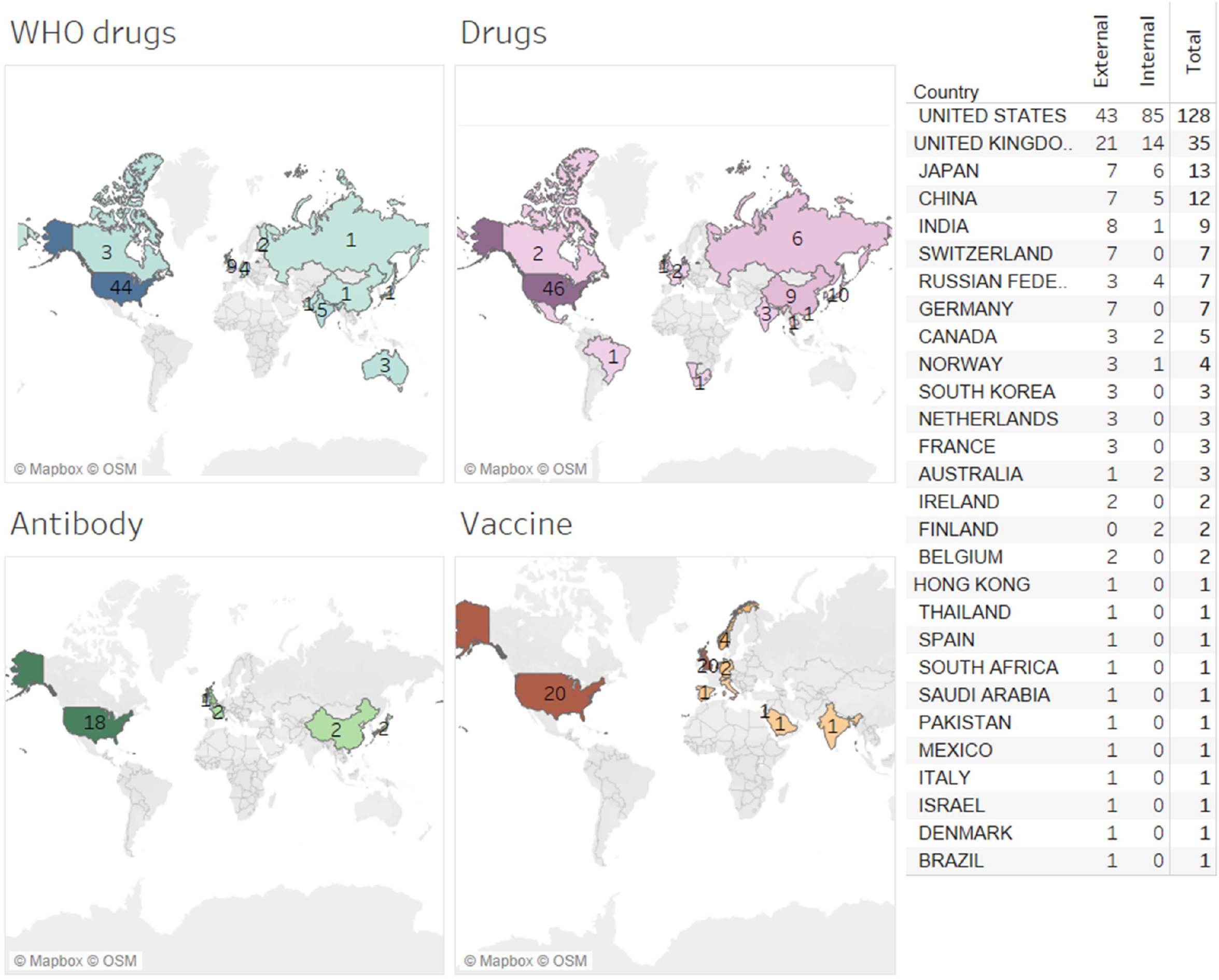
**Agreements on coronavirus infection per country according to the four main groups of therapy. July, 2020. Source: Cortellis**

Regarding the number of documents (Figure 5), the United States and China are the most prominent, with respectively 311 and 174 documents published in the whole period. The majority of these documents (67.8% for China and 58.8% for the United States) were developed without external collaboration, what indicates a high level of research independence of both countries. As for the different groups of drugs and therapies, we also observed a central role of the United States and China. Besides, the maps clearly show that the research on WHO’s drugs is the one that mostly drives the attention and efforts of scientific community, including documents signed by authors from all continents. On the other side, research on coronavirus vaccine does attract or involve a few numbers of countries from restricted regions of the world.

Considering the agreements (Figure 6), the United States once again stands out, being the country with the largest number and share of agreements on coronavirus (almost 50%), while the United Kingdom (and not China) appeared in the second position (with almost 14%). Among the four groups of drugs and therapies, the United States leadership is unbeatable, while European countries show a low level of involvement. It is noteworthy that none of the groups contains agreements authored by countries from all continents, as we observed in Figure 5 for group WHO’s drugs.

Comparing each group in Figures 5 and 6, we noted that countries are found in both maps, but others are replaced or simply disappear, as it is the case of Latin American and African countries that are included in the map of WHO’s drugs in Figure 5 but not in Figure 6. Of course, such different profile may be a consequence of the fact that agreements are signed by two countries only. Nevertheless, it may indicate a gap between countries: in one side the majority of countries that do have technical and human capacities to develop research on coronavirus and in another side just few countries that have these capacities and also the ones needed to develop new forms or products to treat or to prevent the coronavirus.

## CONCLUSIONS AND FINAL REMARKS

The present study aims to investigate whether the drugs indicated by WHO’s Solidarity Project reflect the efforts of the global scientific community related with coronavirus, in other words, in what extent the choice of these drugs is science-based. Such approach, as far as our knowledge, is unique if we consider the lack of similar papers about research on coronavirus in world literature. Besides this characteristic, the present study does develop an innovative strategy search based in MESH terms, which made it possible to retrieve data of 31,815 documents on coronavirus (with no duplication). Such number of documents is indeed high when compared to other similar papers, where the highest number of analyzed documents around 18,000 (38; 39). Hence, considering these two characteristics, we do believe in the originality, robustness and reliability of the data presented in our paper.

Among the general findings, we observed a continuous growth over the decades in three out of the four studied groups of documents (Figure 2), especially the one named as Coronavirus (n. 28,013). In 2020, we observed a big drop in the number of documents in both Gammacoronavirus and Betacoronavirus groups, a clear indication of the great effort of global science to find a quickly response to the human coronavirus responsible for the 2020 epidemic, with a possible shift of researchers from these two groups to beta.

As Coronavirus group encompasses documents of other groups, we considered it as our main source to carry on the following analyzes, including the analysis related to type of research application (Figure 3). With respect to this analysis, we found that the majority of documents are associated with treatment, that correspond to 22.2% of publications included in the Coronavirus group. Contrarily Lou et al (40) found that publications on coronavirus associated to diagnosis and treatment correspond to 10.4%, while those associated to epidemiology are the most frequent, that is 37.2%.

Nevertheless, such discrepancy may be a consequence of the low number of publications on coronavirus analyzed by these authors (n. 183), what made it possible a manual categorization of the publications into different research focuses.

In a second part of our study, that is its core part, we presented findings on documents of Coronavirus group related with drugs and other therapies. We showed that documents in the group WHO’s drugs are much higher than in the other groups (Figure 4 and Table 1), a first indication that WHO’s Solidarity Project was designed upon a robust scientific basis. Nevertheless, it does not mean that all drugs included in this project had the potential to act effectively against betacoronavirs and so in curing COVID-19. In fact, during the development of this study, hydroxychloroquine and the combined drugs lopinavir/ritonavir were inefficient in the COVID-19 clinical trials and so WHO and its partner institutions discontinued their ongoing research. The other two drugs are still in the agenda of global science, but recent results suggest they are as not as much promising in curing or reducing mortality by COVID-19 (41). Despite these disappointments, experts have pointed to many benefits that arose with this big project led by WHO, as stated by Nahid Bhadelia, from the Boston Medical Center “You’re including many different types of subgroups and populations in different parts of the world.” (41)

In fact, WHO’s Project counted on the participation of a wide variety of institutions and countries and so conduct clinical trials all of the world is probably due to the already existing scientific capital and structure in such institutions. It is clear that, among the four groups of drugs and therapies that we investigated, documents related to WHO’s drugs are the ones with the highest involvement of worldwide scientific community (Table 2 and Figure 5). Hence, this is the second sign that WHO’s Solidarity Project was designed upon a scientific basis. A choice of other drugs that had no or very few involvements of institutions and countries would demand a more complex logistic and training and probably it would be more costly.

Our data showed that China and USA share the leadership on coronavirus documents not only in the group of WHO’s drugs but in all other groups of drugs and therapies, being most of these documents with an international collaboration. The strategic position of China and USA as the main leaders of research on coronavirus or COVID-19 is observed in some other studies (40, 42), what corroborates their robust and qualified human workforce as well as their competitive infrastructure. The two nations do also stand out among the top ranked countries in terms of agreements to develop therapies against the coronavirus (Figure 6). Nevertheless, we observed a smaller number of countries signing these agreements, that is, a completely different framework when compared to documents (Figure 5), what may indicate a gap between countries in having technical and human capacities to develop both research on coronavirus or new forms or products to treat or to prevent the coronavirus.

Finally, although our findings indicated the WHO’s choice on the repurposed drugs against COVID-19 was scientific-based in terms of number of published documents and involved institutions, we cannot discard that other drugs may display better performance in both variables. Nevertheless, investigate all available drugs and therapies that are taking part in ongoing research would be a hard (or impossible) task. In any case, we believe that the data shown in this study illustrates that decisions by international organizations, especially in health, may have scientific knowledge as a background and not be merely bureaucratic decisions.

## Data Availability

Data can not be shared because of they were retrieved from secondary sources, which
do not allow the publicity.

## AKNOWLEGMENT

The authors would like to thank the help of doctoral student Stephanie Treiber during the earlier stage of the work. This work has financial support from Capes and CNPq through the Doctoral’s scholarships granted to Andreia Galina and to Deise Sarzi, and the CNPq funding awarded to the research project n. 434.146/2018-8.

## Notes

### Competing Interest Statement

The authors have declared no competing interest.

### Funding Statement

The authors would like to thank the help of the doctoral student Stephanie Treiber
during the earlier stage of the work. This work has financial support from Capes and
CNPq through the Doctoral’s scholarships granted to Andreia Galina and to Deise
Sarzi, and the CNPq funding awarded to the research project n. 434.146/2018-8.

